# COMPARISON OF POSTOPERATIVE PAIN SEVERITY AND ANALGESIC CONSUMPTION WITHIN 24 HOURS BETWEEN PRIMARY AND REPEAT CESAREAN SECTIONS UNDER SPINAL ANESTHESIA: A PROSPECTIVE COHORT STUDY

**DOI:** 10.64898/2026.03.05.26347682

**Authors:** Mequanint Zenebe Bitewlign, Leulayehu Akalu Gemeda, Siryet Tesfaye Delile, Minda Abebe Seife, Mulualem Endeshaw Zeleke, Tesheme Hadush Gebrewahd, Leake Gebregergs Gebreslase, Yohans Tekie Tesfagergse

## Abstract

**Background:** Cesarean section is one of the most commonly performed surgical procedures worldwide and is frequently associated with moderate to severe postoperative pain. While overall pain after cesarean delivery is well described, evidence comparing pain intensity and analgesic use between primary and repeat cesarean sections remains limited.

**Objective:** To compare postoperative pain severity and total analgesic consumption within the first 24 hours among women undergoing primary versus repeat cesarean sections under spinal anesthesia at Tikur Anbessa Specialized Hospital, Addis Ababa, Ethiopia, from January 1 to March 30, 2025.

**Methods:** A prospective cohort study was conducted among 203 women who underwent cesarean delivery under spinal anesthesia. Participants were selected using systematic random sampling and categorized into primary and repeat cesarean groups. Demographic and clinical characteristics were summarized using descriptive statistics. Group comparisons were performed using independent t-tests or Mann–Whitney U tests for continuous variables and Chi-square tests for categorical variables. A p-value < 0.05 was considered statistically significant.

**Results:** Women undergoing repeat cesarean sections experienced significantly higher postoperative pain. During movement, 92.1% of women in the repeat group reported moderate to severe pain compared with 66.7% in the primary group (p < 0.001). At rest, moderate to severe pain occurred in 74.3% of the repeat group versus 52.9% of the primary group (p = 0.002). Pain scores within the first 6 hours were also higher in the repeat group (median NRS 7, IQR 7–8) than in the primary group (median NRS 5, IQR 4–7; p < 0.001). Total analgesic consumption was significantly greater among women in the repeat group (243.3 ± 98.4 mg) compared with the primary group (146.3 ± 82.5 mg; p < 0.001).

**Conclusions:** Repeat cesarean sections are associated with higher early postoperative pain and increased analgesic requirements. These findings support the need for individualized and intensified pain management strategies for women undergoing repeat cesarean delivery.

**Clinical trial number:** Not applicable

## INTRODUCTION

Cesarean section is one of the most common surgeries performed worldwide, with its rates continuing to rise in both developed and developing countries (1). Postoperative pain is a significant concern for women undergoing cesarean section (CS), as it can greatly impact their recovery, mobility, and ability to care for their newborn. During this period, pain is frequently characterized as an unpleasant and upsetting physical and emotional experience (2). As a result of tissue or organ damage that triggers the body’s pain receptors, postoperative pain usually has a nociceptive origin (3). Neuropathic pain can also happen in some situations, especially when nerves are traumatized, stretched, or crushed during surgery (4).

According to international research, acute postoperative pain after CS affects up to 50% of women (1,2). Although the majority of these research emphasize on the prevalence of pain, few evaluate how the primary and repeat CS differ in terms of severity of pain and analgesic requirements. Regarding longer operation times and more complicated surgical challenges such adhesions, some research indicates that women with a history of CS, particularly those with multiple uterine scars, may experience more acute postoperative pain (5,6). Remarkably, other research indicates that women who have repeat cesarean sections sometimes have lower pain ratings and less consumption of opioids than those who have their first CS (7), while these results are still inconclusive. Therefore, it is essential to examine the possible differences in the level of pain severity and analgesics consumed by women having primary and repeat CS in light of these variability’s. Through this, we can improve pain management techniques and make sure that every patient has a safer and more comfortable recovery.

The degree of postoperative pain and the use of analgesics differ depending on whether a woman undergoes a primary or repeat CS, according to several worldwide research. Repeat CS are typically associated with an increased risk of adhesions and other complications that might make surgery more challenging and affect the patient’s pain tolerance (8,9). Nevertheless, there is currently little data that directly compares these two groups’ degrees of postoperative pain. Pain typically starts at the site of the incision in women undergoing a primary CS. Nonetheless, visceral pain is more common in women’s who have had multiple previous cesarean deliveries (1). Scar tissue formation and repeated damage to the same site of anatomy are two factors that increase the chance of chronic or persistent pain with each subsequent surgery(8). In Ethiopia, postoperative pain management is frequently under optimal despite these clinical variations. The lack of knowledge among health care providers about the differences in levels of pain and analgesic need between women undergoing their first cesarean section and those undergoing repeated CS is a significant contributing factor. Lack of knowledge could result in inadequate pain management techniques, which could negatively affect the mother’s health and the surgical procedure as a whole. In order to offer effective individualized pain management, it is essential to fully understand all of these differences.

Therefore, the current study was included patients who are scheduled to undergo primary and repeated cesarean sections to assess the potential difference in postoperative pain severity, and total analgesia consumption among primary and repeated cesarean sections.

## OBJECTIVES

### GENERAL OBJECTIVE

To compare the severity of postoperative pain and analgesic consumption within the first 24 hours among women who underwent a primary cesarean section and those who had a first and second repeat cesarean section under spinal anesthesia at Tikur Anbessa Specialized Hospital, Addis Ababa, Ethiopia, between January 1 and March 30, 2025 G.C.

### SPECIFIC OBJECTIVES

- To compare the severity of postoperative pain, as measured by the Numerical Rating Scale, within the first 24 hours after surgery between women undergoing primary cesarean sections and those undergoing first and second repeat cesarean sections.
- To compare the total amount of analgesic consumption (in milligrams of tramadol or diclofenac equivalent) within the first 24 hours postoperatively between women undergoing primary versus first and second repeat cesarean sections.

## METHODS AND MATERIALS

### Study design

A prospective cohort study was conducted to compare the severity of postoperative pain and the amount of analgesia consumed within the first 24 hours among women who underwent primary cesarean section and those who had their first and second repeat cesarean section. All surgeries were performed under spinal anesthesia at Tikur Anbessa Specialized Hospital (TASH).

### Source of population

All pregnant women who underwent cesarean delivery under anesthesia at TASH.

### Study population

The study population consisted of pregnant women who underwent either elective or emergency cesarean sections under spinal anesthesia at TASH during the study period.

### Study area and period

This study was conducted at TASH, the largest and most well-known teaching and referral hospital in Addis Ababa, Ethiopia. The hospital performs an average of 6 to 12 cesarean deliveries daily, with more than half being elective procedures. The data collection period for this study spanned three months, from January 1 to March 30, 2025.

### Inclusion criteria

Were women aged 18 - 40 years undergoing cesarean section under spinal anesthesia, provided informed consent to participate and ASA physical status II or III.

### Exclusion criteria

Were women’s with history of chronic pain disorders, hearing impairments, failure of spinal anesthesia, Epidural or general anesthesia, presence of severe pregnancy-related complications that were life-threatening and required emergency intervention before the cesarean section, use of additional nerve blocks, multiple pregnancies, IUFD, and three or more previous cesarean scars.

#### Sample size determination

The sample size for this study was determined using the Epi Info statistical calculator, based on data from a previously published comparative studies. In that study, the incidence of severe postoperative pain was reported as 16.7% among primiparous women undergoing a primary cesarean section, and 33.7% among multiparous women undergoing a repeat cesarean section, with findings derived from research conducted in China (1,8). These proportions were used as inputs to ensure adequate statistical power to detect a meaningful difference in pain severity and analgesia consumpssion between the two groups. To verify this estimate, a manual calculation was also performed using the double population proportion formula, assuming: 80% power, 95% confidence interval, and an equal ratio (1:1) between the primary and repeated cesarean section groups. Accordingly, the total calculated sample size was:

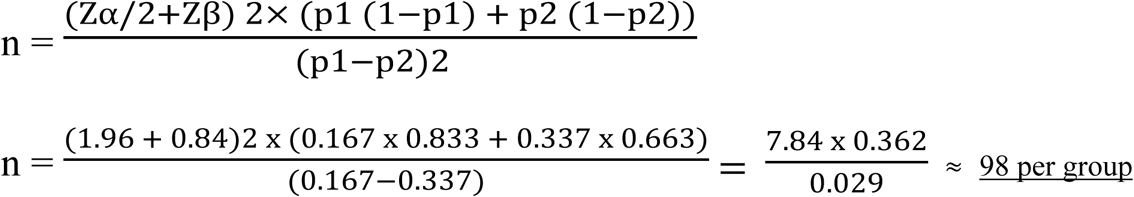

Total sample size = 98 + 98 = 196, including a 5% non-response rate (n = 10), the final calculated sample size was **206.** Where;

- **p1**= 0.167 (proportion of postoperative pain severity in primary C-sections),
- **p2** = 0.337(proportion of postoperative pain severity in repeated C-sections),
- **q** =1-p ; q1 = 1-p1 = 1-0.167 = 0.833 and, q2 = 1-p2 = 1− 0.337 = 0.663,
- **Zα/2** ≈ 1.96 (for a significance level of 0.05), and Zβ ≈ 0.84 (for a power of 0.80)

#### Sampling method

A systematic random sampling method was employed to select eligible pregnant women from the daily surgical schedule. The first participant was randomly selected using a lottery method from the day’s scheduled patients posted on the operation board. Subsequently, data were collected from every second eligible patient until the required sample size was reached within the study period.

**FIGURE 1:**
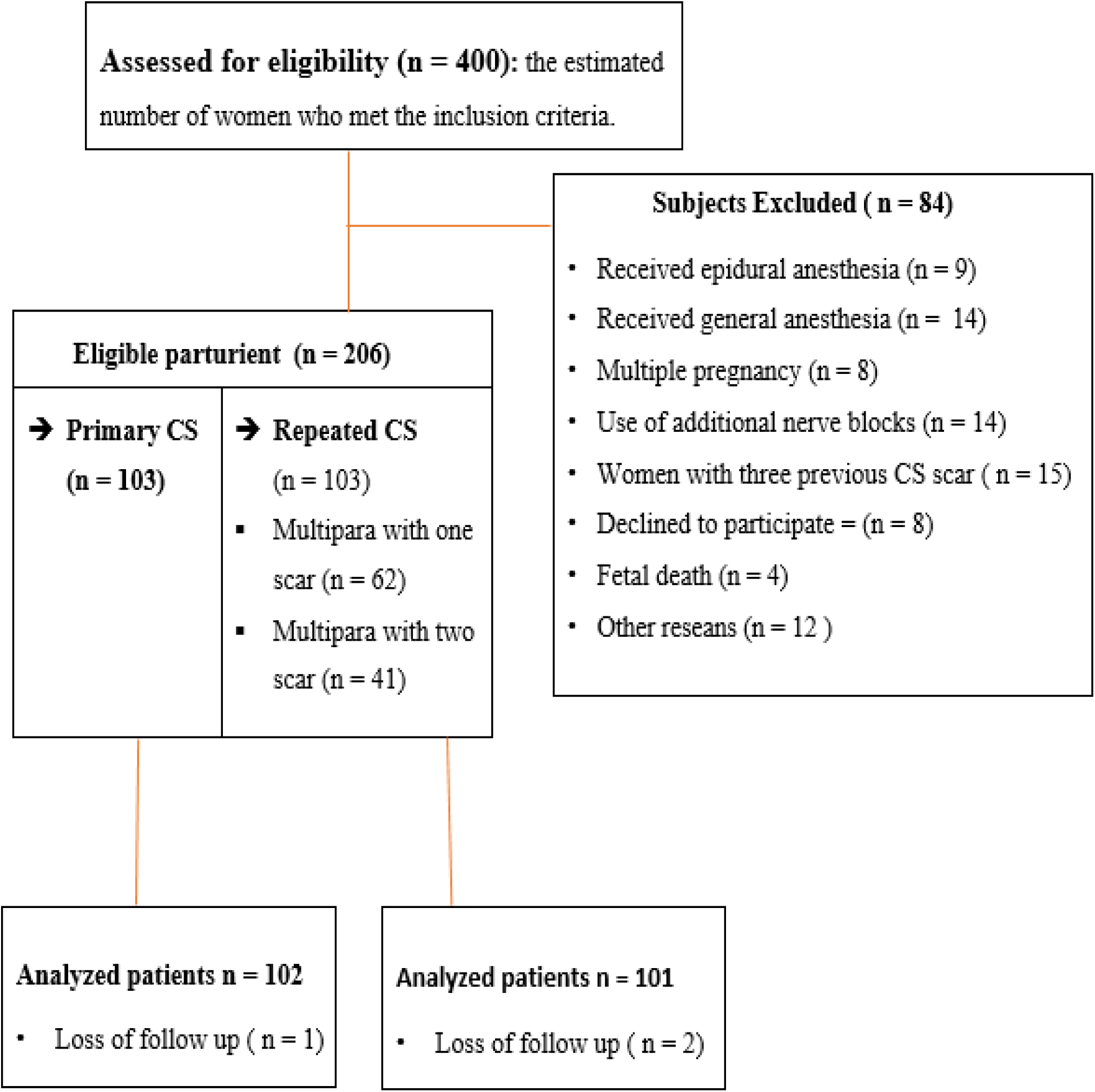
FLOW DIAGRAM ILLUSTRATING THE SELECTION PROCESS OF STUDY PARTICIPANTS.

### Study variables

#### Dependent variables

- Postoperative pain severity and
- Total post-operative analgesia consumption

### Independent variables

#### Maternal Socio demographic Factors

Age, Body Mass Index (BMI), ASA physical status classification, Educational status and marital status.

#### Preoperative obstetric characteristics

Presence of preoperative pain, Preoperative stress, Parity, Type of cesarean section (primary, first and second repeat), Indication for cesarean delivery and History of previous other surgeries

#### Intraoperative anesthetic and surgical factors

Duration of surgery, Intraoperative use of analgesics, Type of surgical incision, Type of local anesthesia used and Dose of local anesthetic and any adjuvant agents used

#### Post-operative pain characteristics, analgesia used and recovery experiences

Type and amount of postoperative analgesics administered (including tramado (IV), diclofenac (intramuscular injections and suppositories)), Type of Pain, Character or nature of Pain, Pain experienced at rest and during movement (coughing).

### Operational definitions

▪ **Cesarean Section:** A surgical procedure used to deliver one or more babies through an incision in the mother’s abdomen and uterus, performed under anesthesia (29).
▪ **Primary Cesarean Section:** The first surgical delivery performed on a woman under spinal anesthesia (29).
▪ **Repeated Cesarean Sections:** Refers to the first and second repeat surgical deliveries performed after a previous cesarean section under spinal anesthesia (29).
▪ **Postoperative Pain (POP):** Any pain reported by the patient following surgery, indicated by a pain score (NRS) greater than zero (28).
▪ **Postoperative inadequate analgesia:** Defined as a NRS score of ≥ 4, representing moderate to severe pain (8).
▪ **Severity of postoperative pain:** Defined as a NRS score of ≥ 7, representing severe pain (2,8).
▪ **Total Postoperative Analgesia Consumption:** The total amount of analgesic drugs (in milligrams) administered within the first 24 hours following the patient’s admission to the post-anesthesia care unit (7,28,30).
▪ **Numerical Rating Scale:** A simple, widely used method of pain assessment tool where the patient identifies their current pain level on a scale of 0 to 10. Pain intensity was assessed using the NRS at 1, 6, 12, and 24 hours postoperatively (2,17,28).

### Data collection procedures and tools

Data collection was carried out at TASH by three trained data collectors through chart reviews and structured questionnaires available in both English and Amharic. The questionnaire included sections on socio-demographic characteristics, preoperative obstetric information, intraoperative details, and postoperative pain characteristics, patient pain responses at rest and during movement, recovery experiences, and follow-up information. A Numerical Rating Scale (NRS) was used to measure the intensity of postoperative pain.

During the preoperative visit, the first data collector (an anesthetist) explained the study’s objectives, potential risks and benefits, and confidentiality procedures to eligible participants. Informed consent was obtained from all participating mothers. In the operating room, patients were connected to standard ASA monitors. Spinal anesthesia was administered using 0.5% Bupivacaine at the L3–L4 interspace in a sitting position, after confirming free flow of cerebrospinal fluid without barbotage. Following administration, the anesthetist assessed the block level: autonomic block with alcohol swabs, sensory block with a pinprick test, and motor block using the Bromage scale (target score of 3). Only patients who achieved an adequate sensory level (T5-T6) and a successful spinal block were included in the study. Intraoperative data were documented accordingly.

At the conclusion of surgery, patients were transferred to the Post-Anesthesia Care Unit (PACU), where immediate postoperative pain scores during the first hour were assessed using the NRS, both at rest and during movement (during cough). The patient’s bed number and assigned ward were documented before transfer from the PACU, and a formal handover was provided to the ward nurse. In the postoperative ward, the other two data collectors (nurses) assessed and recorded both incisional and visceral pain scores using the static NRS (at rest during quiet breathing) and dynamic NRS (after voluntary coughing). The women were asked to report their pain intensity using NRS at the, 6th, 12th, and 24th hours post-surgery (1,2,21,28). These scores were recorded from the patients’ charts, and the total analgesic consumption within the first 24 hours was also documented for each patient.

### Data processing, analysis and data quality management

To ensure the quality and reliability of the data, the questionnaire was pre-tested on 5% of the calculated sample size. Prior to analysis, thorough data cleaning and verification were performed to ensure data integrity. The data were then coded and entered into Microsoft Excel, and subsequently exported to SPSS version 27 for statistical analysis.

The normality of continuous variables was assessed using the Shapiro – Wilk test. Descriptive statistics were used to summarize the data, with findings presented in tables and figures. For continuous variables: If data were normally distributed, group comparisons were made using the independent t-test and results were expressed as mean ± standard deviation. If data were not normally distributed, the Mann–Whitney U test was used, and results were presented as median and interquartile range. For categorical variables: The Chi-square test was employed to assess differences between groups, with results expressed as frequencies (n) and percentages (%).

A p-value < 0.05 was considered to indicate statistical significance throughout the analysis.

### Ethical consideration

The research was carried out after obtained ethical approval from the Institutional Review Board of Addis Ababa University, School of Medicine, and Department of Anesthesia. After participants were fully made aware of the significance of the study, process, and objective, all the participants provided written consent after being fully informed. There was neither a material nor financial incentive for participating. All participants were fully made aware of their right to withdraw at any time from the study without risking any penalty.

## RESULT

### Group (CS type)

#### SOCIO-DEMOGRAPHIC CHARACTERISTICS

203 women who had cesarean sections (CS) at TASH in 2025 were included in the study. The average age in the primary CS group was 26.89 years (± 4.74), while women in the repeated CS group was mean age of 30.86 years (± 4.22; p < 0.001). There were no statistically significant differences between the two groups in terms of educational level, marital status, weight, height, BMI, or ethnicity (p > 0.05), as shown in Table 1 above.

**TABLE 1:** DISTRIBUTION OF SOCIO - DEMOGRAPHIC CHARACTERISTICS OF PATIENT WHO UNDERWENT CAESAREAN SECTION (CS) AT TASH, 2025 (n = 203).

| Variable | Category | Group (CS type) |  | P value |
| --- | --- | --- | --- | --- |
|  |  | Primary<br>n = 102 | Repeated<br>n = 101 |  |
| **Age** | Years ( $\bar{X} \pm SD$) | 26.89 ± 4.74 | 30.86 ± 4.22 | < 0.001* |
| **Weight** | In kilogram | 66.95 ± 9.97 | 66.6±10.69 | P = 0.811 |
| **Height** | In meters | 1.61 ± 0. 065 | 1.61 ± .058 | P = 0.924 |
| **Educational level** | No formal education n (%) | 7(6.9) | 10(9.9) | P = 0.327 |
|  | Read and write n (%) | 38(37.3) | 42(41.6) |  |
|  | Secondary school n (%) | 28(27.5) | 31(30.7) |  |
|  | Degree and above n (%) | 29(28.4) | 18(17.8) |  |
| **Marital status** | Single n (%) | 0(0.0) | 1(1.0) | P = 0.368 |
|  | Married n (%) | 101(99) | 100(99) |  |
|  | Windowed n (%) | 1(1.0) | 0(0.0) |  |
| **BMI** | Underweight (<18.5) n (%) | 0(0.0) | 1(1.0) | P = 0.743 |
|  | Normal weight (18.5-24.9) n (%) | 47(46.1) | 48(47.5) |  |
|  | Over weight (25-29.9) n (%) | 43(42.2) | 39(38.6) |  |
|  | Obesity (30-39.9) n (%) | 12(11.8) | 13(12.9) |  |
| **Ethnicity** | Amhara n (%) | 38(37.3) | 31(30.7) | P = 0.487 |
|  | Oromo n (%) | 21(20.6) | 23(22.8) |  |
|  | Tigray n (%) | 4(3.9) | 3(3.0) |  |
|  | Gurage n (%) | 22(21.6) | 32(31.7) |  |
|  | Slte n (%) | 10(9.8) | 9(8.9) |  |
|  | Others n (%) | 7(6.9) | 3(3.0) |  |
| **Residence** | Urban n (%) | 101(99) | 101(100) | P = 0.318 |
|  | Rural n (%) | 1(1.0) | 0(0.0) |  |

**Note:** Values are presented as number (%), analyzed using the Chi-square test; and as mean ± standard deviation (SD), analyzed using the independent t-test. BMI refers to Body Mass Index. A p-value < 0.05 was considered statistically significant.

#### PREOPERATIVE OBSTETRIC CHARACTERISTICS

A significantly greater proportion of women in the primary CS group were classified as ASA physical status II (94.1%) compared to the repeated CS group (85.1%). Parity and the number of children were significantly higher among women in the repeated CS group (p < 0.001 for both), a notably higher proportion of women in the repeated CS group reported no preoperative pain (85.1%) compared to those in the primary CS group (56.9%; p < 0.001). Emergency CS were more frequently performed in the primary group (65.7%), whereas the repeated CS group had a higher proportion of elective procedures (82.2%; p < 0.001), as seen in table 2 above.

**TABLE 2:** PREOPERATIVE CHARACTERISTICS OF THE PARTURIENT WHO UNDERWENT CAESAREAN SECTION AT TASH, 2025 (n =203).

| Variable | Category | Group |  | P value |
| --- | --- | --- | --- | --- |
|  |  | Primary n = 102 | Repeated n = 101 |  |
| ASA | II | 96(94.1%) | 86(85.1%) | P = 0.036 |
|  | III | 6(5.9%) | 15(14.9%) |  |
| Parity | 1 times | 63(61.8%) | 2(2%) | < 0.001** |
|  | 2 times | 16(15.7%) | 38(37.6%) |  |
|  | 3 times | 18(17.6%) | 46(45.5%) |  |
|  | > or = 4 times | 5(4.9%) | 15(14.9%) |  |
| Number of children's | 1 | 68(66.7%) | 3(3%) | < 0.001** |
|  | 2 | 17(16.7%) | 54(53.5%) |  |
|  | > or = 3 | 17(16.7%) | 44(43.6%) |  |
| Preoperative pain (using the NRS ) | No pain | 58(56.9%) | 86(85.1%) | < 0.001 |
|  | Mild pain | 13(12.7%) | 6(5.9%) |  |
|  | Moderate pain | 23(22.5%) | 9(8.9%) |  |
|  | Severe pain | 8(7.8%) | 0(0.0%) |  |
| Reason for CS | Elective | 35(34.3%) | 83(82.2%) | < 0.001 |
|  | Emergency | 67(65.7%) | 18(17.8%) |  |
| Preoperative stress | Low n (%) | 39(38.2%) | 39(38.6%) | P = 0.937 |
|  | Medium n (%) | 46(45.1%) | 47(46.5%) |  |
|  | High n (%) | 17(16.7%) | 15(14.9%) |  |

**Note:** Values are presented as number (%) and analyzed using the Chi-square test. ASA refers to the American Society of Anesthesiologists physical status classification. A p-value < 0.05 was considered statistically significant.

### INTRAOPERATIVE ANESTHETIC AND SURGICAL - RELATED FACTORS

The duration of surgery was significantly longer in the repeated cesarean section group compared the primary CS group (p < 0.001). This extended time is likely due to the presence of adhesions. The type of surgical incision, did not differ between the groups (P = 0.314). There were no statistically significant differences between the two groups in terms of intraoperative analgesia and the use of adjuvant medications (p > 0.05, as shown in Table 3 above.

**TABLE 3:** INTRAOPERATIVE ANESTHETIC AND SURGICAL - RELATED FACTORS OF THE PARTURIENT WHO UNDERWENT CAESAREAN SECTION AT TASH, 2025 (n =203).

| Variable | Category | Group |  | P value |
| --- | --- | --- | --- | --- |
|  |  | Primary n=102 | Repeated n=101 |  |
| Dose of bupivacaine given | 10mg | 24(23.5%) | 16(15.8%) | P = 0.334 |
|  | 12.5mg | 63(61.8%) | 66(65.3%) |  |
|  | 15mg | 15(14.7%) | 19(18.8%) |  |
| Adjuvant added | Morphine | 2(2.0%) | 5(5.0%) | P = 0.147 |
|  | None | 94(92.2%) | 90(89.1%) |  |
|  | Dexamethasone | 3(2.9%) | 0(0.0%) |  |
|  | Fentanyl | 3(2.9%) | 6(5.9%) |  |
| Duration of surgery | In minutes ( $\bar{X} \pm \text{SD}$ ) | 41.7 ± 4.1 | 51.7 ± 5.4 | P < 0.001** |
| Type of incision | Classical [midline] | 0(0.0%) | 1(1.0%) | P = 0.314 |
|  | Pfannenstiel | 102(100%) | 100(99.0%) |  |
| Intraoperative analgesia used | None | 95(93.1%) | 98(97.0%) | P = 0.436 |
|  | Tramadol | 5(4.9%) | 2(2.0%) |  |
|  | Fentanyl | 2(2.0%) | 1(1.0%) |  |

**Note:** Values are presented as number (%) and were analyzed using the Chi-square test; mean ± standard deviation (SD) values were analyzed using the independent t-test. A p-value < 0.05 was considered statistically significant.

#### COMPARISON OF POSTOPERATIVE PAIN SEVERITY AMONG PRIMARY AND REPEATED CESAREAN SECTION

**TABLE 4:**
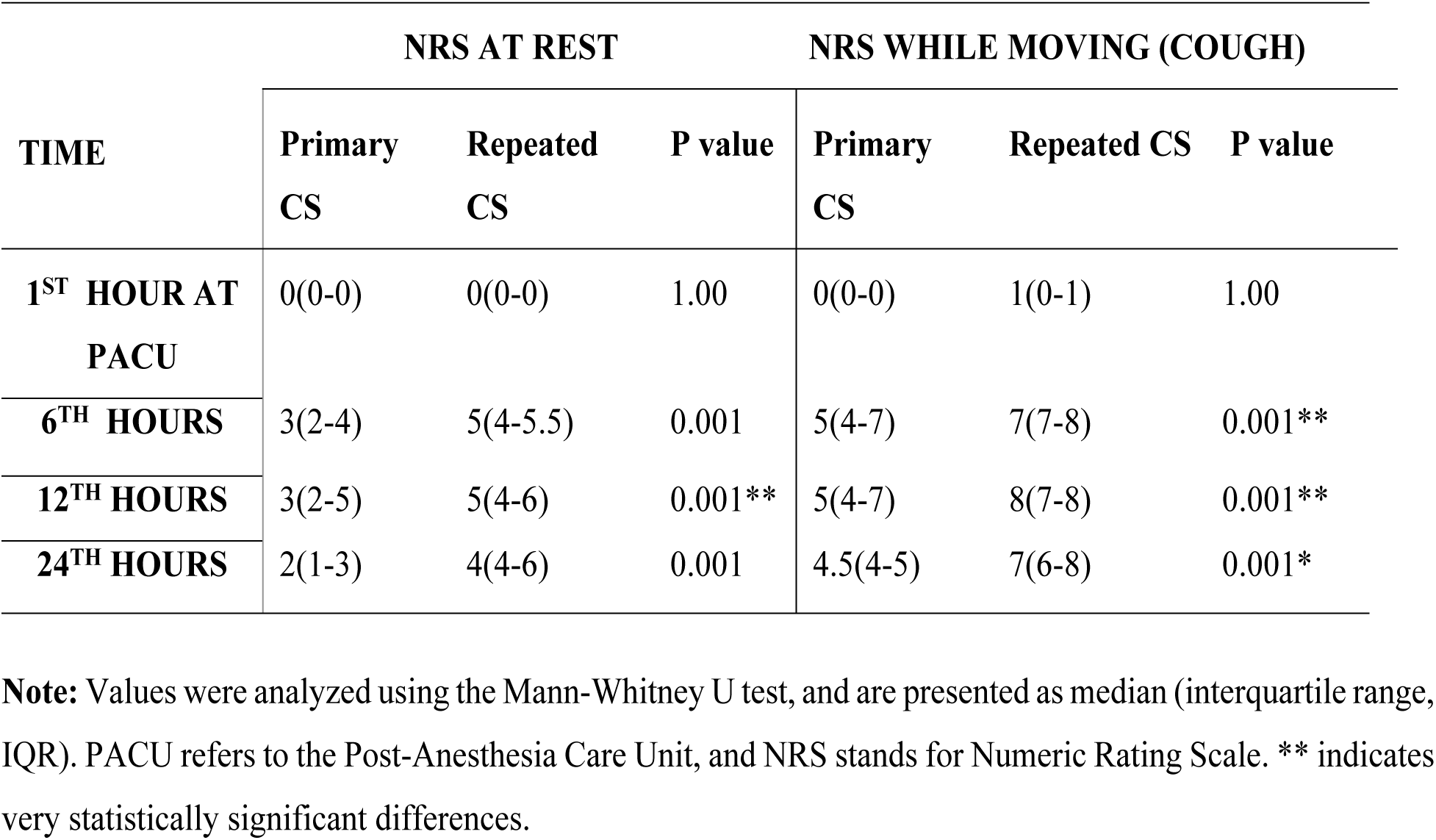
Comparison of postoperative pain severity by NRS at rest and while moving at TASH, 2025 (n=203)

**Table 5:** Incidence of moderate to severe postoperative pain at rest and while moving (during cough) for mothers who underwent caesarean section at TASH, 2025 (n=203)

| Group |  |  |  |  |  |  |
| --- | --- | --- | --- | --- | --- | --- |
| Variable | Primary n = 102 |  | Repeated n = 101 |  | RR 95% CI | P value |
|  | Moderate to Severe POP n (%) |  | Moderate to Severe POP n (%) |  |  |  |
| *Pain intensity* | NO | YES | NO | YES |  |  |
| **6<sup>th</sup> hrs.** |  |  |  |  |  |  |
| At rest | 48(47.1) | 54 (52.9) | 26(25.7) | 75(74.3) | 1.82(1.2 -2.7) | P = 0.002 |
| At moving | 34(33.3) | 68 (66.7) | 8 (7.9) | 93 (92.1) | 3.03(1.6 -5.7) | < 0.001*** |
| **12<sup>th</sup> hrs.** |  |  |  |  |  |  |
| At rest | 55(53.9) | 47 (46.1) | 27 (26.7) | 74 (73.3) | 1.97(1.4-2.8) | < 0.001** |
| At moving | 36(35.3) | 66 (64.7) | 17(16.8) | 84(83.2) | 1.75(1.2-2.6) |  |
| **24<sup>th</sup> hrs.** |  |  |  |  |  |  |
| At rest | 62(60.8) | 40(39.2) | 38(37.6) | 63(62.4) | 1.6(1.2-2.2) | P = 0.001* |
| At moving | 53(52) | 49(48) | 31 (30.7) | 70 (69.3) | 1.7(1.1-2.3) | P = 0.002 |

**Note:** Values are presented as frequency (proportion) were analyzed using the Chi-square test. Abbreviations: RR – Relative Risk, POP – Postoperative Pain, hrs. – Hours, CI – Confidence Interval. \*\*\* indicates highly statistically significant differences.

**FIGURE 2:**
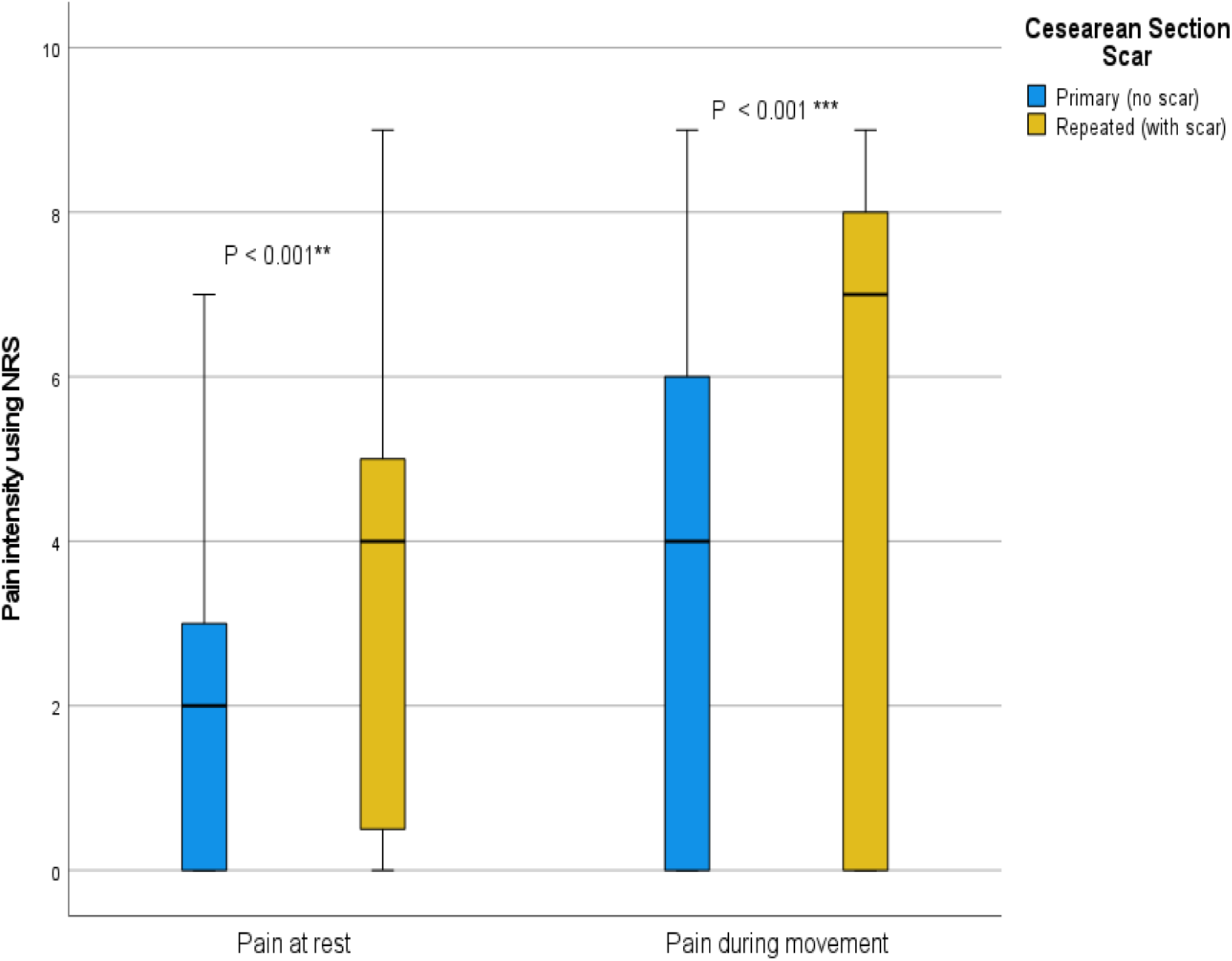
BOX PLOT SHOWING THE PAIN SCORES AT REST AND DURING MOVEMENT (COUGHING) WITH IN THE FIRST 24 HOURS POSTOPERATIVELY. *** indicates highly statistically significant differences.

At the 6-hour mark, a significant difference in pain severity during movement was observed: 66.7% of patients in the primary CS group experienced moderate to severe pain, compared to 92.1% in the repeated CS group (p < 0.001). At rest during the 12-hour mark, 73.3% of repeated CS patients reported moderate to severe pain (p < 0.001). Movement-induced pain at 12 hours was especially pronounced in repeated CS patients, with 83.2% experiencing moderate to severe pain compared to just 64.7% in the primary group (p < 0.001). By 24 hours after surgery, overall pain intensity had declined, which may be attributed to the natural physiological reduction of inflammatory mediators involved in the pain response.

The type of pain also varied markedly between the two groups. In the primary CS group, incisional pain alone was predominant (77.5%), whereas in the repeated CS group, the majority (78.2%) experienced a combination of incisional and visceral pain (p < 0.001) **(see fig. 3 below**). This difference may be due to increased internal manipulation, adhesions, or scarring associated with previous surgeries.

**FIGURE 3:**
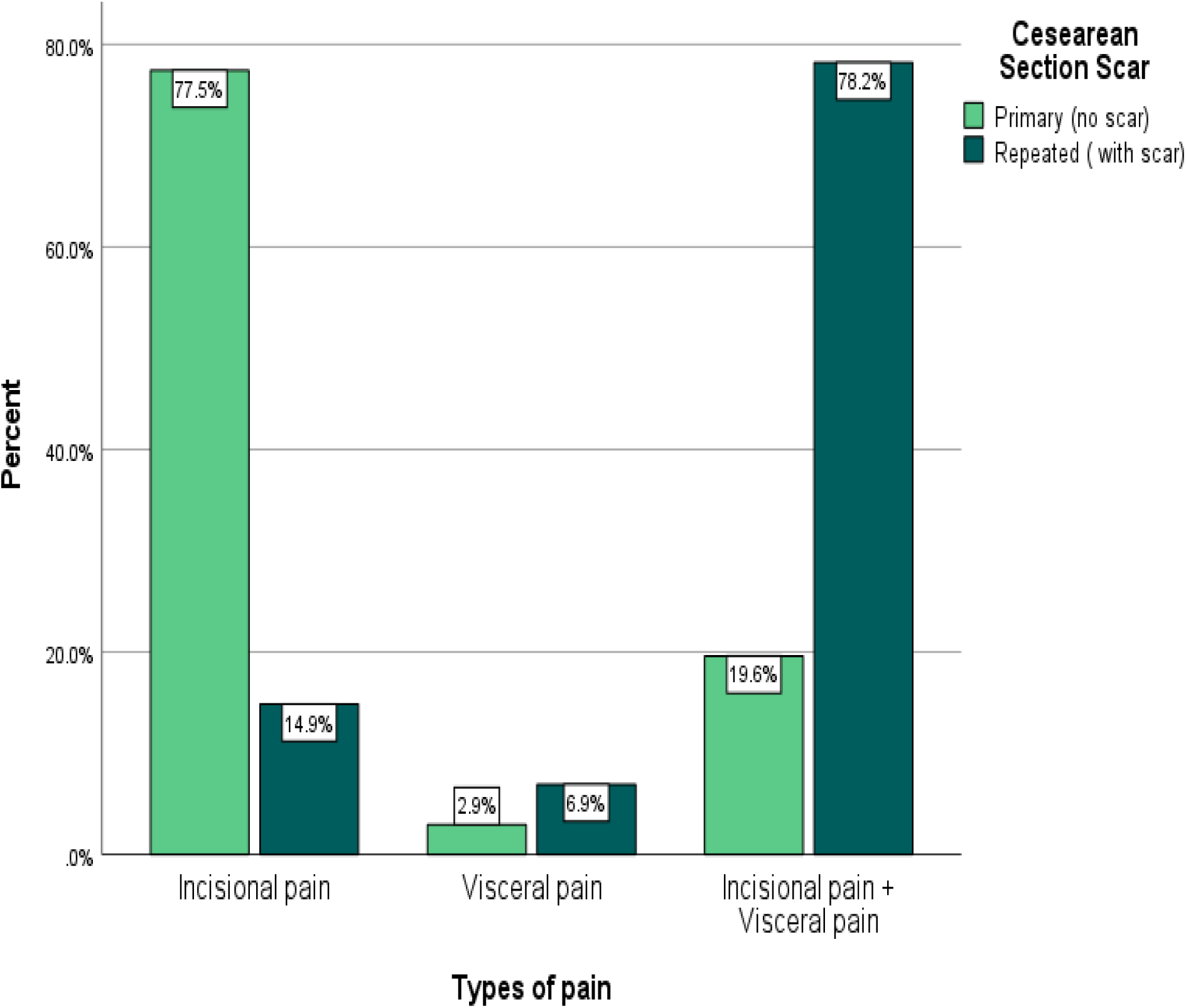
BAR GRAPH ILLUSTRATING THE TYPES OF POSTOPERATIVE PAIN AMONG WOMEN UNDERGOING PRIMARY AND REPEAT CESAREAN SECTIONS.

#### COMPARISION OF POST-OPERATIVE ANALGESIA CONSUMPTIONS, PAIN CHARACTERISTICS, AND RECOVERY EXPERIENCES AMONG PRIMARY AND REPEATED CESEREAN SECTION

**TABLE 6:**
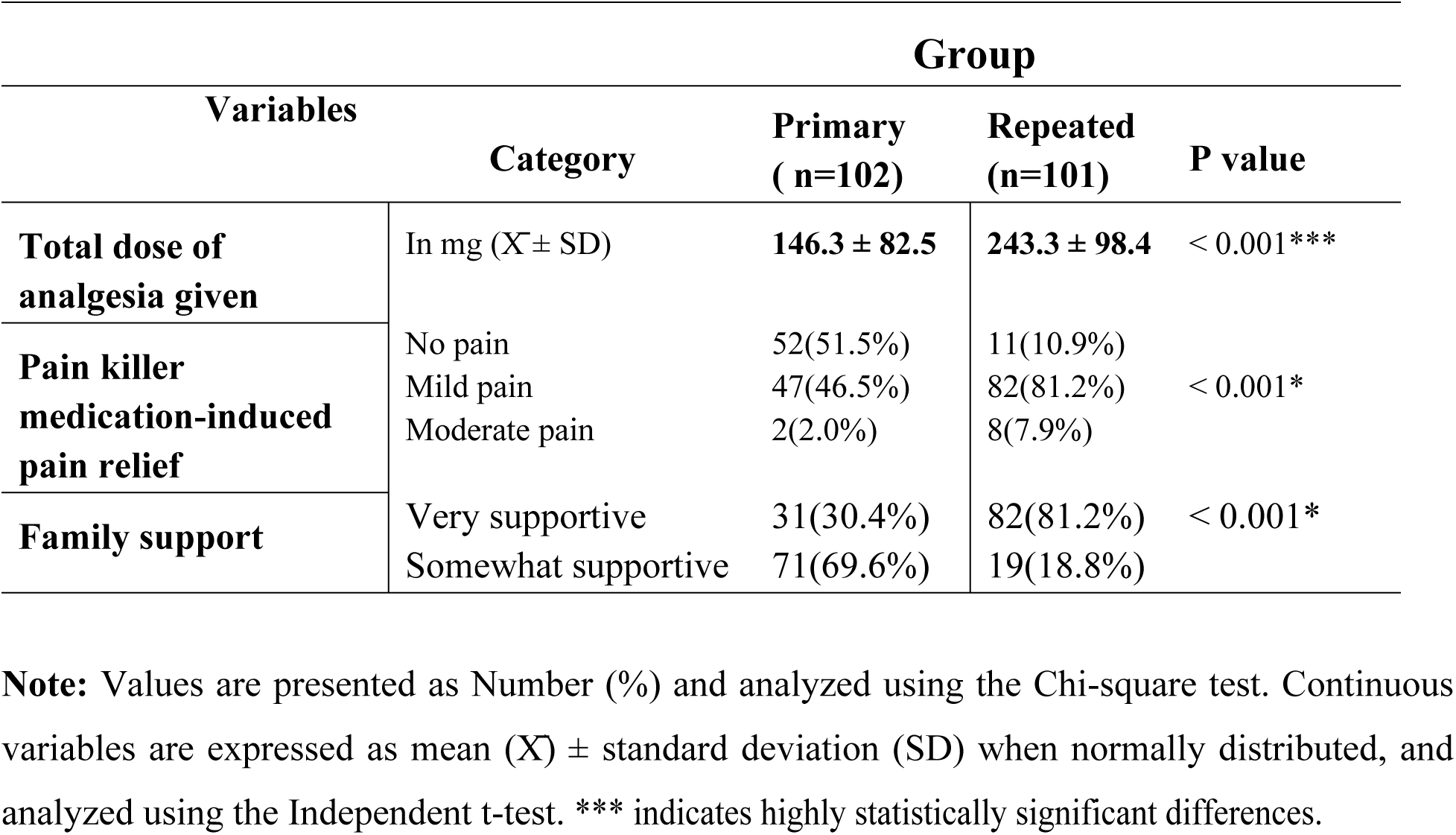
COMPARISON OF POST-OPERATIVE PAIN CHARACTERISTICS, ANALGESIA CONSUMPTION AND RECOVERY EXPERIENCES WITHIN 24 HOURS AMONG PARTURIENT WHO UNDERWENT CAESAREAN SECTION AT TASH, 2025 (n =203)

|  |  | Group |  |  |
| --- | --- | --- | --- | --- |
| Variables | Category | Primary<br>( n=102) | Repeated<br>(n=101) | P value |
| **Total dose of analgesia given** | In mg ($\bar{X} \pm SD$) | **146.3 ± 82.5** | **243.3 ± 98.4** | < 0.001*** |
| **Pain killer medication-induced pain relief** | No pain | 52(51.5%) | 11(10.9%) | < 0.001* |
|  | Mild pain | 47(46.5%) | 82(81.2%) |  |
|  | Moderate pain | 2(2.0%) | 8(7.9%) |  |
| **Family support** | Very supportive | 31(30.4%) | 82(81.2%) | < 0.001* |
|  | Somewhat supportive | 71(69.6%) | 19(18.8%) |  |

**Note:** Values are presented as Number (%) and analyzed using the Chi-square test. Continuous variables are expressed as mean ( $\bar{X}$ ) ± standard deviation (SD) when normally distributed, and analyzed using the Independent t-test. \*\*\* indicates highly statistically significant differences.

## DISCUSSION

This study demonstrates that women undergoing repeated cesarean sections experience significantly greater postoperative pain and require higher doses of analgesics compared to those undergoing a primary cesarean delivery. In particular, 92.1% of women in the group that had a repeat cesarean section experienced moderate to severe pain during movement, while only 66.7% of women in the primary cesarean group did the same. Women who have had previous cesarean sections had a significantly higher pain burden, as indicated by the computed relative risk of 3.03 (95% CI: 1.6–5.7). When evaluating postoperative pain, individual aspects including the existence of surgical scars should be considered since they may contribute to this enhanced pain experience.

In repeat CS, the median NRS score for post-operative pain severity was 7 (IQR: 7–8), while in the primary cesarean group, it was 5 (IQR: 4–7) (p < 0.001). The pain scores of both groups of patients were similar in the first hour after surgery, however there were differences by the sixth hour. At 6-, 12-, and 24-hours post-surgery, women who had repeated caesarean sections experienced higher pain severity during rest and exercise, particularly during coughing (p < 0.001). In the first 24 postoperative hours, the repeat cesarean group took significantly more analgesics than the primary cesarean sections, with an average of 243.3 mg (±98.4) against 146.3 mg (±82.5) (p < 0.001). This large range highlights the significance of personalizing postoperative pain management techniques, particularly for individuals undergoing repeat procedures.

Our findings align with Getahun et al.’s (2025) study, which found that 91.1% of women in the repeat cesarean group experienced moderate to severe pain while coughing or moving, compared to 52.4% in the primary group. Pain at rest was more common in repeat cesareans (74.3% in this research and 87.5% in Getahun’s), compared to primary cesarean patients (52.9% and 35.7%, respectively), with a relative risk of 2.45 (95% CI: 1.98-3.02; p < 0.001) (28). Studies consistently reveal that repeated cesarean sections cause more severe pain following surgery, both at rest and during movement. This could be due to scar tissue, adhesions, or more complex surgical dissection, all of which can cause tissue trauma and slow wound healing.

Our results are similarly in line with a study conducted in China by Yang et al., which found that multiparous women experienced a higher rate of inadequate analgesia (24.0%) than primiparous women undergoing cesarean section (16.7%) (8). But according to our research, a significantly greater percentage (92.1%) of women in the group that had multiple cesarean sections reported having moderate to severe pain, with a relative risk of 3.03. The application of patient-controlled analgesia (PCA) in Yang et al.’s study could account for this discrepancy, and which could have affected pain management and analgesic consumption. Additionally, the increased surgical complexity in repeated cesareans such as scarring, adhesions, and internal manipulation likely contributed to the higher pain levels observed in our cohort.

Contrary to our findings, a study by Gürbüz et al. conducted in Turkey assessed postoperative pain within the first six hours and reported a mean pain score of 5.56 ± 1.31 on the Visual Analog Scale, reported with no significant difference in pain levels based on the number of previous cesarean sections (24). However, our findings clearly reveal that women who have had many cesarean deliveries feel considerably more postoperative pain than those who have had a primary cesarean section. Pain intensity in the repeat cesarean section increased from a median NRS score of 7 (IQR: 7-8) at 6 hours post-surgery to 8 (IQR: 7-8) by 12 hours. At the same time period, however, the primary cesarean section reported a median NRS pain score of 5 (IQR: 4-7). These inequalities can be attributed to variations in pain measurement methodologies, timing of assessment, therapeutic interventions, and individual or cultural, social differences in pain perception.

This study’s findings are consistent with those of Duan et al. in China, who followed both primiparas and multipara women undergoing multiple cesarean sections for 48 hours after surgery. The study indicated that women who had several cesarean sections had higher rates of postoperative pain, with a reported relative risk of 3.56 (95% CI: 1.05-12.04) (1). Demelash et al. found a link between multiple cesarean sections and higher postoperative pain (AOR: 2.3; 95% CI: 1.1-5.0), suggesting that previous procedures may contribute to post-operative pain severity (13). These findings support our findings and emphasize the need for personalized pain management measures for women enduring repeat cesarean procedures.

Study conducted by Duan et al. suggested that only 22.7% of women with repeated cesarean sections reported visceral pain, but incisional pain was more common in primary cesarean section (38.3%) (1). Our study found that women who had multiple cesarean sections (78.2%) experienced both incisional and visceral pain, while those in the primary cesarean section mostly reported incisional pain (77.5%). Our study used the Numerical Rating Scale, while Duan’s used the Visual Analog Scale, potentially explaining the variations in pain measurement methods. Variations may also be explained by characteristics such as surgical experience, number of previous procedures, and pain threshold. Increased scarring and adhesions from repeated operations are likely to contribute to more complex and extensive pain.

The current study found that cumulative postoperative analgesic consumption was higher in the repeat cesarean group than in the primary cesarean group. The finding of this study concurs with that of Getahun et al.’s prospective cohort study, which found that women in the repeat cesarean group required earlier analgesics and also had higher cumulative consumption of analgesics. The following are some reasons why this similarity may occur: same study design, increased tissue damage, adhesions from prior surgery, and longer operative times associated with repeated cesareans, which may all lead to more severe postoperative pain and greater doses of analgesics (28).

Our study revealed that women undergoing repeated cesarean section needed significantly higher doses of analgesics in the first 24 hours after surgery (p < 0.001) than those who had primary cesarean sections. In contrast to our findings, Chao A. et al. in the United States, who reported lower opioid consumption and no significant differences in postoperative pain ratings (7). This gap could be due to variations in patient characteristics, clinical assessment, or pain treatment approaches. One possible reason for our study higher pain and need for analgesics is that they had more severe adhesions or changed pain sensitivity due to previous operations. These findings underscore the importance of adapting pain treatment options to each patient’s individual needs and clinical status.

As compared to the United States study by Holmes et al. where no difference was observed in opioid use or pain scores between primary and multiple cesarean section (22), our study showed significantly greater consumption of analgesic and postoperative pain severity among women who were exposed to repeated cesarean sections. This could be explained by the effect of surgical trauma, the number of previous surgeries, or variations in pain management types and strategies, population variability, and the current study’s prospective cohort methodology. These disparities underscore the challenges of controlling pain following repeated cesarean sections.

## LIMITATION OF STUDY

Frist, the research was limited in external validity and its capacity to be generalized to more diverse or larger healthcare settings due to its single-institution design.

Second, only the first 24 hours postoperatively were used in order to quantify severity of pain and consumption of analgesic, making it unable to examine effects longer-term such as chronic postoperative pain.

## STRENGTH OF STUDY

A validated measure of pain assessment instrument (Numerical Rating Scale) was also utilized within the study at various rest and movement time points.

Statistical power and stability of the findings were enhanced by the sample size, which was quite large in relation to the previous research.

## CONCLUSION

This study concludes that compared to women who have the primary cesarean section, women who undergo multiple cesarean sections are much more likely to experience moderate to severe postoperative pain and need more analgesics. These results imply that when developing postoperative pain management policies, individual differences like surgical scars should be carefully taken into account.

## RECOMMENDATION

We recommend that medical and anesthesia staff consider surgical scarring and history of previous cesarean sections when planning management of postoperative pain. In particular, in women having repeat cesarean sections, pain management has to be individualized.

In a better effort to understand their individual pain control needs, more than three women with previous cesarean sections must be enrolled in future studies, beyond the first 24 hours post operation, long-term recovery and pain need to be evaluated and multicenter randomized trials conducted to ascertain that the results are more universally applicable.

## Data Availability

The data cannot be shared publicly because they contain sensitive patient information. Access to the data can be requested from the Institutional Review Board of Addis Ababa University (contact) for researchers who meet the criteria for confidential data access.

## LIST OF ACRONMYS AND ABBREVIATIONS

ASA: American society of anesthesiology
BMI: Body mass index
CS: Cesarean section
CD: Cesarean delivery
CI: Confidence interval
ETB: Ethiopian Birr
G.C: Gregorian colanders
IQR: Interquartile range
IM: Intramuscularly
IV: Intravenously
NRS: Numerical rating scale
OR: Odd ratio
PI: Principal investigator
POP: Postoperative pain
PACU: Post-anesthesia care unit
PCA: Patient-Controlled Analgesia
SPSS: Statistical package for the social sciences
SD: Standard deviation
WHO: World Health Organization
RCT: Randomized controlled trial
RR: Relative risk
TASH: Tikur Anbessa Specialized Hospital
USA: United States of America
VAS: Visual analog scale

## DECLARATIONS

### Ethics approval and consent to participate

Ethical approval was obtained from the Institutional Review Board of Addis Ababa University, College of Health Sciences ***(Protocol No. Anes 02/2024/25).*** Official permission to conduct the study was also secured from Tikur Anbessa Specialized Hospital.

Written informed consent was obtained from all participants prior to enrollment. Confidentiality was maintained by removing personal identifiers from the dataset and securely storing all collected information. All study procedures were conducted in accordance with the Declaration of Helsinki.

### Consent for publication

Not applicable.

### Availability of Data and Materials

The datasets analyzed during the current study are available from the corresponding author upon reasonable request.

### Authors’ Contributions

MZB conceived and designed the study, selected the study title, wrote the proposal, performed statistical analysis, interpreted the findings, drafted the manuscript, and approved the final version for publication. LAG, STD, and MAS contributed to the conceptual design, data entry, and critically revised the final manuscript for important intellectual content. MEZ, THG, LGG, and YTT assisted in synthesizing information, writing, and editing the final manuscript. All authors have read and approved the submitted manuscript and agree to be accountable for all aspects of the work.

## Acknowledgements

The author would like to thank Addis Ababa University for academic support and the staff of Tikur Anbessa Specialized Hospital for their cooperation during data collection. Special appreciation is extended to all study participants.

